# Exome sequencing identifies *ABCA7* as an important gene for familial AD cases from Eastern India

**DOI:** 10.1101/2024.08.30.24312765

**Authors:** Dipanwita Sadhukhan, Adreesh Mukherjee, Bidisha Bhattacharyya, Smriti Mishra, Tapas Kumar Banerjee, Gautam Das, Uma Sinharoy, Subhra Prakash Hui, Soma Gupta, Atanu Biswas, Arindam Biswas

## Abstract

**Introduction:** AD is the most complex disorder leading to dementia worldwide. Despite the disease burden among Indians the mutation spectrum in our subcontinent is not well examined.

**Methods:** To identify probable causal variants for AD, in a total of 29 clinically diagnosed, majorly familial and young onset AD cases from Eastern India, whole exome analysis in Illumina NovoSeq. 6000 sequencing platform was performed as per standard methodology. Next, the coding variants were prioritised based on the literature and our bioinformatic analyses for genotype-to-phenotype correlation.

**Results:** A total of 25 missense variants and 4 nonsense variants in 17 genes among 23 AD cases were identified as probable damaging ones in our study cohort. Amongst all, *ABCA7* account for the maximum number of pathogenic variants (5/29). A lowering in the age of onset was observed for mutation carriers and cases belonging to the posterior cortical atrophy (PCA) subgroup. Our further comparative analyses suggested that variants in APP metabolism pathway genes are more common in PCA, frontal AD, and young-onset multi-domain amnestic phenotypes than dysexecutive and typical AD.

**Conclusion:** Our study suggests that whole exome sequencing among Indian AD patients holds the potential to identify the most common gene as well as clinically relevant new causal variants which may highlight new insight into disease mechanism through future research.

## INTRODUCTION

Alzheimer’s disease (AD) is the most common progressive neurodegenerative disorder, with varying degrees of manifestation between individuals of the same ethnicity. Among Indians, the prevalence rate of dementia is 7.4% (around 8.8 million individuals) over the age of 60 years (Lee et al., 2023) of which AD alone or in combination with vascular pathology constitutes the majority. However, in contrast to European Ancestry, little is known about the Indian genetic architecture conferring AD susceptibility. The maximum number of reports are on the genetic association of AD with ApoE4 (Bharath et al., 2010, Shankarappa et al., 2017). However, an earlier Whole exome sequencing (WES) has been successful in the identification of pathogenic AD-associated novel variants among Indians (Syama et al., 2018). Therefore, considering complex disease mechanisms, little knowledge of AD genetics, and highly heritable polygenic AD susceptibility (h2=0.58-0.79) (Karlsson et al., 2022), the present study aimed to adopt a WES strategy to highlight the genes probably associated with Familial Alzheimer’s Diseases (FAD) from eastern India.

## MATERIALS AND METHODS

### Study subjects

Based on radiological findings and standard diagnostic criteria for AD by National Institute on Aging and Alzheimer’s Association (NIA-AA) Workgroup criteria (Jack et al., 2024) here a total of 29 AD subjects with either positive family history or younger age of onset were recruited by Cognitive Neurologists and Neuropsychologists from the Bangur Institute of Neurosciences and National Neurosciences Centre Calcutta, Kolkata, India. Among them, 22 had a younger age of onset (< 65 years), and 24 had a positive family history of AD or dementia.

### Whole exome sequencing

To perform WES, genomic DNA was extracted using the QIAamp DNA Blood kit (QIAGEN, Germany) followed by targeted gene capture using a custom capture kit. Paired-end sequencing was performed with 2×100/2×150 chemistry on Illumina NovoSeq 6000 sequencing platform. Reads were assembled and aligned to reference sequences based on NCBI RefSeq transcripts and human genome build GRCh37/UCSC hg19.

### In-silico analyses

The variation calling, annotation, and prioritization were performed to identify the variants of interest and interpreted in the context of a single clinically relevant transcript. All variants identified in known 36 genes for AD and other dementia-related genes/loci were manually evaluated based on documented literature, online database and bioinformatics prediction. The nonsynonymous variants that showed damaging effects by more than 60% of tools used (13/21) or more would be considered probable damaging and reported here.

## Results

### Demographic details of the study cohort

To understand the genetic basis of AD in the Eastern Indian AD population, we performed a pilot WES in a total of 29 AD patients with cognitive impairment for an average of more than 1.5 years at the time of presentation. The mean age at onset is 55.03±11.59 years and age ranges from 35 to 74 years. The male-to-female ratio is 0.6111. An except for 5 cases, all of them have a positive family history.

### Identification of genetic variants

Among the plethora of variants identified, here we report a total of 25 missense variants and 4 nonsense variants with high pathogenicity in 17 genes among 23 AD cases representing ∼79.31% (23/29) cases as carriers.

It includes majorly five missense mutations in *ABCA7* (distributed throughout the protein), three in *SORL1* (a novel and two reported), two novel variants like Phe383Val and Leu258Ser in *PSEN1* (in TM6, TM7) and the pathogenic Val717Ile in *APP* gene. Apart from these, dementia and PD-related genes also appeared to carry pathogenic variants among AD patients (Table 1a). The Ser15ProfsTer39 of *EIF4G1*, Trp218Ter of *GBA*, Phe7LeufsTer80 of *UNC13B*, and Lys484Ter of *VPS13C* variants account for the nonsense variant identified here.

**Table 1a:** Description of genetic variants with a higher pathogenicity prediction score by *in-silico* tools.

| Gene Name | cDNA change | Amino acid change | Protein domain | dbSNP ID | MAF (gnomAD v4.0.0) | Functional Prediction: Damaging (Total) | Comments |
| --- | --- | --- | --- | --- | --- | --- | --- |
| *APP* | c.2149G>A | Val717Ile | Intracellular domain | rs63750264 | 0.000002401 | 17 (21) | Amyloid precursor protein |
| *PSEN1* | c.1156T>C | Phe386Val | TM7 | Novel | Not available | 20 (21) | Proteolytic subunit of gamma-secretase, role in processing in APP |
|  | c.785T>C | Leu258Ser | TM6 | Novel | Not available | 20 (21) |  |
| *ABCA7* | c.1547G>C | Arg516Pro | Extracellular loop 1 | rs377020137 | 0.00008299 | 15 (21) | Transporter involved in cholesterol metabolism and clearance of Aβ |
|  | c.2974A>G | Ile992Val | NBD motif | rs574186698 | 0.00002544 | 15 (21) |  |
|  | c.4590G>A | Met1530Ile | 2<sup>nd</sup> TM helix | rs747309721 | 0.000001593 | 13 (21) |  |
|  | c.4606G>A | Val1536Ile | Extracellular loop | rs371168746 | 6.843E-07 | 15 (21) |  |
|  | c.5548G>A | Glu1850Lys | NBD motif | rs530609497 | 6.911E-07 | 17 (21) |  |
| *SORL1* | c.1A>G | Met1Val | VPS1 domain | Novel | Not available | 15 (20) | Role in APP processing and lysosomal targeting of Aβ |
|  | c.808G>A | Glu270Lys | VPS10 domain | rs117260992 | 0.01757 | 16 (20) |  |
|  | c.3643C>T | Arg1215Trp | LDLR class A domain | rs776459268 | 0.00001054 | 17 (21) |  |
| *CCNF* | c.62G>A | Arg21Gln | F-box | rs200973445 | 0.000005576 | 16 (21) | Associated with FTD/ALS |
|  | c.1217G>A | Arg406Gln | Cyclin N-terminal | rs146438273 | 0.00197 | 17 (21) |  |
| *DNAJC13* | c.3191A>G | Asp1064Gly | N-terminal to J domain | Not available | 0.000002478 | 18 (21) | Associated with PD |
|  | c.3887A>G | Glu1296Gly | N-terminal to J domain | rs61748101 | 0.006254 | 18 (20) |  |
|  | c.4546C>T | Arg1516Cys | C-terminal to J domain | rs79734612 | 0.0008753 | 16 (20) |  |
|  | c.5018A>G | Tyr1673Cys | C-terminal to J domain | rs79953286 | 0.05256 | 15 (20) |  |
| *EIF4G1* | c.42_43del | Ser15ProfsTer39 | N-terminal domain | Novel | Not available | Not applicable | Associated with dementia |
| *GBA* | c.653G>A | Trp218Ter | Glyco-hydro_30 domain | rs867929413 | 0.000002738 | Not applicable | Associated with dementia |
| *CD2AP* | c.62G>A | Arg21Gln | 1<sup>st</sup> SHC domain | rs559978848 | 0.0001289 | 13 (21) | Regulates Aβ generation by neuron-specific polarization of Aβ in dendritic early endosome |
| *EPHA4* | c.1150G>C | Val384Leu | Fibronectin type III domain | Not available | Not available | 14 (21) | Regulates spine morphology and plasticity in the brain |
| *LRRK2* | c.5237C>A | Ala1746Asp | ROC-COR domain | Novel | Not available | 18 (21) | Processing in APP |
| *NOS3* | c.466G>A | Glu156Lys |  | rs141456642 | 0.0003607 | 19 (21) | Associated with AD |
| *PLA2G6* | c.1123G>A | Glu375Lys | ANK7 domain | Not available | 0.0000012 | 17 (21) | Associated with neurodegeneration |
| *SPAST* | c.1532A>C | Glu511Ala | AAA-ATPase domain | CD103676 | 0.000001593 | 15 (21) | Regulation of microtubule dynamics |
| *TREM2* | c.155G>C | Arg52Pro | Ig-V set domain | CM1413005 | Not available | 17 (21) | Immunoglobulin superfamily receptor binds lipids and Aβ |
| *UNC13B* | c.21del | Phe7LeufsTer80 | C2A domain | rs372094156 |  | Not applicable | Involved in synaptic vesicle exocytosis |
| *VPS13C* | c.1450A>T | Lys484Ter | VPS13-alpha | Novel | Not available | Not applicable | Associated with PD |
|  | c.7445G>A | Arg2482His | Putative WD40 domain | rs115481870 | 0.009195 | 16 (20) |  |

Twenty-one online software were used including SIFT, PolyPhen2-HDIV, PolyPhen2-HVAR, MutationTaster, MutationAssessor, FATHMM, PROVEAN, VEST4, MetaSVM, MetaLR, M-CAP, CADD, DANN, fathmm-MKL, Eigen, GenoCanyon, fitCons, GERP++, phyloP, phastCons and SiPhy.
Damaging (Total) is the ratio of online software that predicted the variant as damaging among the 21 online software. cDNA: complementary deoxyribonucleic acid; dbSNP: dbSNP137 (https://www.ncbi.nlm.nih.gov/snp/); MAF gnomAD: the minor allele frequency of variants in the genome aggregation database (gnomAD v4.0.0).

In contrast, missense variants mentioned in Table 1b (Arg566Cys in *EIF4G1*, Pro455Leu & Ala2075Glu in *ABCA7* and Arg202Gln in *GBA* genes) showed less damaging effects upon analyses.

**Table 1b:** Clinical features of AD cases harboring potential pathogenic/likely pathogenic variants.

| Sl. No. | Lab ID | Age at onset (in ranges) | Family History | Gender | ApoE status | Disease duration (yrs.) | Gene 1 | Amino acid change/Nucleotide variants | Gene 2 | Amino acid change | Clinical presentation | Additional features |
| --- | --- | --- | --- | --- | --- | --- | --- | --- | --- | --- | --- | --- |
| 1. | DEM247 | 45-50 | Positive | M | e3e4 | 1.5 | *APP* | Val717Ile | *SOLRI* | Glu270Lys | Young-onset MDA | Parkinsonism |
| 2. | DEM334 | 55-60 | Positive | M | e3e3 | 2 | *APP* | Val717Ile | - | - | Young-onset MDA | Nil |
| 3. | DEM307 | 45-50 | Negative | F | e3e4 | 1.5 | *PSEN1* | Leu258Ser | - | - | PCA | Pyramidal sign |
| 4. | DEM323 | 35-40 | Positive | F | e3e3 | 2.5 | *PSEN1* | Phe386Val | - | - | PCA | Nil |
| 5. | DEM262 | 40-45 | Negative | F | e3e3 | 2 | *ABCA7* | Arg516Pro | - | - | Frontal AD | Parkinsonism |
| 6. | DEM268 | 45-50 | Positive | F | e2e3 | 2.5 | *ABCA7* | Ile992Val | - | - | PCA | Nil |
| 7. | DEM264 | 65-70 | Positive | M | e3e4 | 3 | *ABCA7* | Met1530Ile | *PLA2G6* | Glu375Lys | Typical AD | Nil |
| 8. | DEM239 | 45-50 | Positive | F | e3e4 | 2 | *ABCA7* | Val1536Ile | - | - | Frontal AD | Parkinsonism |
| 9. | DEM290 | 65-70 | Positive | F | e3e4 | 3 | *ABCA7* | Glu1850Lys | - | - | Typical AD | Mild Parkinsonism |
| 10. | DEM226 | 70-75 | Positive | F | e3e4 | 2.5 | *SORL1* | Met1Val | - | - | Typical AD | Nil |
| 11. | DEM284 | 50-55 | Positive | M | e3e3 | 2.5 | *SORL1* | Arg1215Trp | *DNAJC13* | Glu1296Gly | PCA | Nil |
| 12. | DEM283 | 35-40 | Negative | F | e3e4 | 2 | *CCNF* | Arg21Gln | *LRRK2* | Ala1746Asp | PCA | Nil |
| 13. | DEM233 | 40-45 | Positive | M | e3e4 | 2 | *CCNF* | Arg406Gln | *DNAJC13* | Asp1064Gly | Frontal AD | Nil |
| 14. | DEM332 | 75-80 | Positive | F | e3e4 | 2 | *DNAJC13* | Arg1516Cys | - | - | Typical AD | Nil |
| 15. | DEM244 | 45-50 | Negative | F | e3e4 | 1 | *DNAJC13* | Tyr1673Cys | - | - | Young-onset MDA | Nil |
| 16. | DEM305 | 50-55 | Positive | M | e3e4 | 2 | *GBA* | Trp218Ter | - | - | Typical AD | Parkinsonism |
| 17. | DEM291 | 65-7 | Positive | M | e4e4 | 2 | *EIF4G1* | Ser15ProfsTer39 | - | - | Typical AD | Nil |
| 18. | DEM235 | 60-65 | Positive | F | e4e4 | 1.5 | *EPHA4* | Val384Leu | - | - | Typical AD | Nil |
| 19. | DEM328 | 55-60 | Negative | F | e3e3 | 2 | *CD2AP* | Arg21Gln | - | - | Typical AD | Nil |
| 20. | DEM315 | 45-50 | Positive | F | e3e3 | 1.5 | *NOS3* | Glu156Lys | *SPAST* | Glu511Ala | PCA | Nil |
| 21. | DEM329 | 60-65 | Positive | F | e3e3 | 2 | *TREM2* | Arg52Pro | - | - | Frontal AD | Nil |
| 22. | DEM311 | 60-65 | Positive | F | e3e4 | 1 | *VPS13C* | Lys484Ter | *VPS13C* | Arg2482His | PCA | Nil |
| 23. | DEM287 | 40-45 | Positive | F | e3e4 | 3 | *UNC13B* | Phe7LeufsTer80 | - | - | Dysexecutive | Nil |
| 24. | DEM227 | 65-70 | Positive | F | e3e3 | 3 | *EIF4G1* | Arg566Cys | - | - | Typical AD | Nil |
| 25. | DEM229 | 60-65 | Positive | F | e3e4 | 1.5 | *ABCA7* | c.5848-5C>T | - | - | PCA | Nil |
| 26. | DEM240 | 70-75 | Positive | F | e3e4 | 1.5 | *NOTCH3* | Leu33del | - | - | Typical AD | Nil |
| 27. | DEM269 | 65-70 | Positive | M | e3e4 | 3 | *ABCA7* | Pro455Leu | - | - | Typical AD | Nil |
| 28. | DEM295 | 40-45 | Positive | M | e3e4 | 2 | *ABCA7* | Ala2075Glu | - | - | PCA | Nil |
| 29. | DEM335 | 60-65 | Positive | M | e3e4 | 2 | *GBA* | Arg202Gln | *NOTCH3* | Arg2145Gln | Dysexecutive | Nil |

PCA: Posterior Cortical Atrophy; MDA: Multidomain Amnesic; AD: Alzheimer's Disease

### Phenotypic variability and relation to genotype

To explain the genotype-to-phenotype relation, it is noteworthy to mention that sixteen patients had a monogenic mutation and 7 cases were digenic/double mutants for these genes.

Typical AD, Posterior Cortical Atrophy (PCA), and Frontal variant of AD appeared as common clinical features followed by young-onset multi-domain amnestic and dysexecutive syndrome (Table 1b) in our study cohort.

Our preliminary comparisons of the age of onset between the groups showed that it is a) lowest for PCA and highest for typical AD (P-value=0.0423 and 0.0001297 respectively) and b) lower among the mutation carriers than non-carriers (53±11.21 vs 62.83±10.40; P=0.08438) (Table 2a).

**Table 2a:**
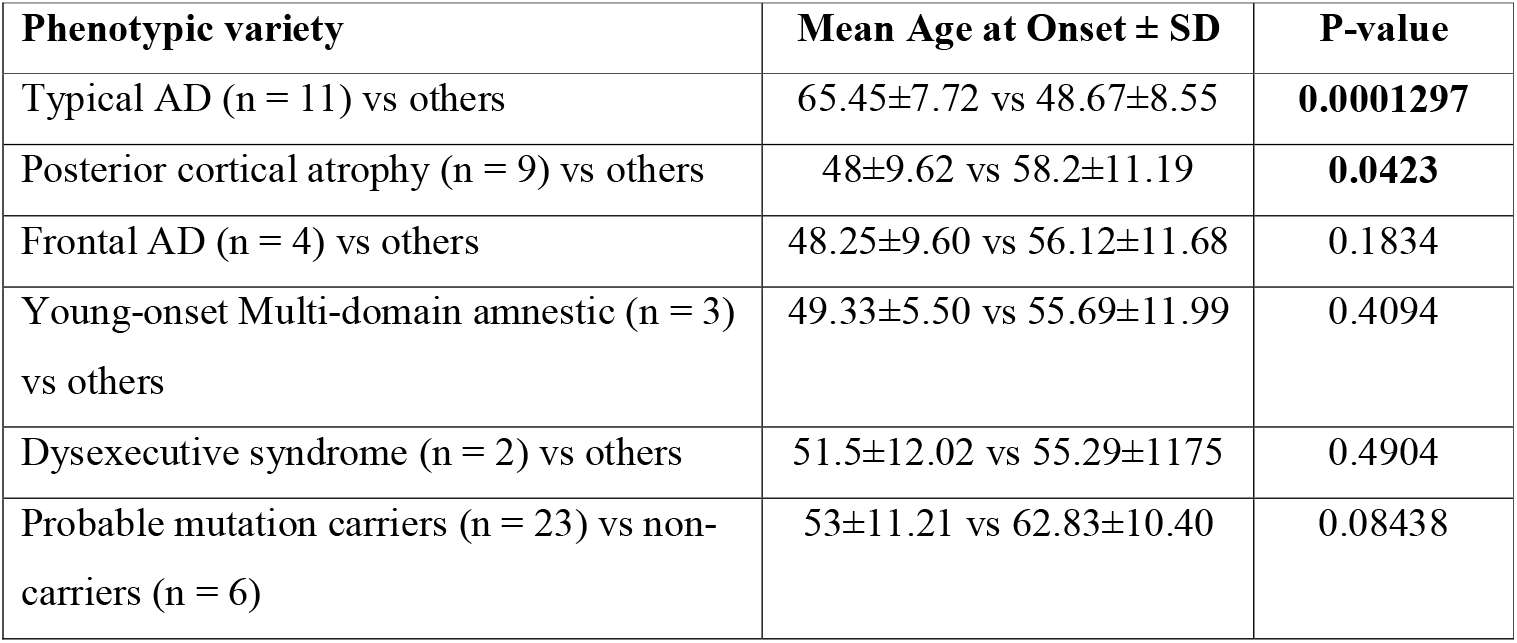
Comparison of Age at onset for different subgroups of AD.

Our further inter-group analysis showed that the genetic variants in the APP metabolism (for PCA 78%, for frontal variant 75%, for young-onset multi-domain amnestic 67%, and 36% for typical AD) and non-APP metabolism pathway (100% for the dysexecutive syndrome, 64% for typical AD, 33% for young-onset multi-domain amnestic, 25% for frontal variant, and 22% for PCA) are present across phenotypes in different frequencies (Table 2b). Therefore, the categorization of genes linked with AD according to metabolic pathway suggests that alterations in genes of the APP metabolism pathway are likely to result in PCA, frontal AD and young-onset multi-domain amnestic, while, non-APP metabolism pathway involvement may show dysexecutive, and typical AD.

**Table 2b:** Comparison between phenotypic variability with mean age at onset and APP metabolism pathway.

| Phenotypic variety | Pathway involved | Mean Age at Onset ± SD | P-value |
| --- | --- | --- | --- |
| Typical AD (n = 11) | APP metabolism (4; 36%) | 67.75±2.87 | 0.6335 |
| | Non-APP metabolism (7; 64%) | 64.14±9.47 | |
| Posterior cortical atrophy (n = 9) | APP metabolism (7; 78%) | 46.28±9.30 | 0.5565 |
| | Non-APP metabolism (2; 22%) | 54±11.31 | |
| Young-onset Multi-domain amnesic (n = 3) | APP metabolism (2; 67%) | 52±4.24 | 0.6667 |
|  | Non-APP metabolism (1; 33%) | 44 |  |
| Frontal variant (n = 4) | APP metabolism (3; 75%) | 49.67±11.23 | 1.00 |
|  | Non-APP metabolism (1; 25%) | 44 |  |
| Dysexecutive syndrome (n = 2) | Non-APP metabolism (2; 100%) | 51.5±12.02 | Not applicable |

The APOE4 load is quite high here; 18 individuals are found with the *APOE4* allele in heterozygous condition and two were homozygous for the same. The *E4* allele is present irrespective of the different AD variants.

## DISCUSSION

Therefore, in the present study, after screening 29 AD patients from Eastern India we have identified several probable pathogenic variants in a total of 17 genes majorly in *ABCA7, SORL1, APP* and *PSEN1*.

The London variant in the *APP* gene that we identified here too, is known as the mutational hotspot for autosomal dominant AD. An increased Aβ_1–42_/Aβ_1–40_ leading to more pathological protein accumulation is the underlying mechanism for it. On the other hand, *PSEN1* variants located in its transmembrane domains, TM6 and TM7 respectively are novel and uncharacterised. However, since these variants share the same domains with the two major catalytic aspartic acids (Asp275 and Asp385) which are important for their endoproteolytic activity and γ-secretase activity, here we speculate for a deficiency in protein’s function as their patho-mechanism towards early-onset AD. Amongst all the genes, *ABCA7* was identified to carry the maximum number of variants which are clustered in Extracellular loop1 and NBD motif followed by 2nd transmembrane helix. A variability in the age of onset for *ABCA7* carriers was observed in our study. Several factors like the location of variants, *APOE* genotype, and the presence of additional variants in other AD or PD-related genes could be reasons for such variable clinical manifestations. Unlike a previous study examining *ABCA7* variants in AD cases with memory and non-memory cognitive impairment, here we found severe memory impairment in all carriers (Campbell et al., 2022). In line with growing evidence from candidate and GWAS studies, our study has also identified *SORL1* as an important gene conferring risk to AD in familial cases. Our intergroup analysis also highlighted the contribution of non-APP pathway genes across different AD varieties even in a very small sample size.

An earlier study from India on AD-exome analysis already identified variants in genes involved in APP metabolism like ours, but the profile is different concerning location in protein and frequency (Syama et al., 2018). However, considering these two studies, we can say APP-metabolism genes contribute significantly to familial AD among Indians. Moreover, our Indian data regarding the influence of *APOE* genotype on phenotypic heterogeneity is in accordance with global data.

Therefore, in conclusion, genes related to Aβ formation and clearance can be termed as major players in Indian familial AD cases and future exome analyses in larger cohorts have the potential to identify new variants in common genes which can provide new disease insight through model study.

## Acknowledgement

The authors thank the patients who participated in the study.

## Funding

Supported by grants from the Department of Science & Technology, Govt. of India, under the Cognitive Science Research Initiative Programme (DST/CSRI-P/2017/22), and Department of Biotechnology, Ministry of Science & Technology, Govt. of India (BT/NIDAN/01/05/2018).

## Credit authorship contribution statement

DS and AB were responsible for the concept, study design, experimental work, data analysis and manuscript preparation.

AM, AB, GD, US, and TKB were responsible for the clinical diagnosis, concept, study design and manuscript preparation.

BB and SM were responsible for data acquisition and manuscript preparation. SPH and SG were responsible for manuscript preparation.

All authors read the draft, provided their inputs and agreed on the final version of the manuscript.

## Conflict of Interest

There is no conflict of interest.

## Ethics statement

All procedures performed in studies involving human participants were in accordance with the ethical standards of the IPGME&R, Kolkata, and National Neurosciences Centre Calcutta, Kolkata, India. The Ethics Committees (Institutional Ethics Committee, IPGME&R, Kolkata and National Neurosciences Centre Calcutta, Kolkata) of the abovementioned Institutes approved the study protocol. Informed consent was taken as per guidelines of the Indian Council of Medical Research, National Ethical Guidelines for Biomedical and Health Research involving human participants, India.

## Informed consent

Informed consent from all the participants was received before clinical data and sample collection.

## Data Availability Statement

The data described in this study are available from the corresponding author upon reasonable request.

## Notes

### Competing Interest Statement

The authors have declared no competing interest.

